# Health-related quality of life in Long COVID: Mapping the condition-specific C19-YRSm measure onto the EQ-5D-5L

**DOI:** 10.1101/2024.08.11.24311809

**Authors:** Adam B. Smith, Darren C. Greenwood, Paul Williams, Joseph Kwon, Stavros Petrou, Mike Horton, Thomas Osborne, Ruairidh Milne, LOCOMOTION Consortium, Manoj Sivan

## Abstract

**Background:** Long Covid (LC) is a clinical syndrome of persistent, fluctuating symptoms subsequent to COVID-19 infection with a prevalence global estimate of many millions of cases. LC has significant detrimental effects on health-related quality of life (HRQoL), activities of daily living (ADL), and work productivity. Condition-specific patient-reported outcome measures (PROMs), such as the modified Covid-19 Yorkshire Rehabilitation Scale (C19-YRSm), have been developed to capture the impact of LC. However, these do not provide health utility data required for cost-utility analyses of LC interventions. The aim of this study was therefore to derive a mapping algorithm for the C19-YRSm to enable health utilities to be generated from this PROM.

**Methods:** Data were collected from a large study evaluating LC services in the UK. A total of 1434 people with LC had completed both the C19-YRSm and the EQ-5D-5L on the same day. The EQ-5D-5L responses were then converted to EQ-5D-3L utility scores. Correlation and linear regression analyses were applied to determine items from the C19-YRSm and covariates for inclusion in the algorithm. Model fit, mean differences across the range of EQ-5D-3L scores (−0.59 to 1), and Bland-Altman plots were used to evaluate the algorithm. Responsiveness (standardised response mean; SRM) of the mapped utilities was also investigated on a subset of participants with repeat assessments (N=85).

**Results:** There was a strong level of association between 8 items and 2 domains on the C19-YRSm with the EQ-5D single-item dimensions. These related to joint pain, muscle pain, anxiety, depression, walking/moving around, personal care, ADL, and social role, as well as Overall Health and Other Symptoms. Model fit was good (R^2^ = 0.7). The mean difference between the actual and mapped scores was < 0.10 for the range from 0 to 1 indicating a good degree of targeting for positive values of the EQ-5D-3L. The SRM for the mapped EQ-5D-3L health utilities (based on the C19-YRSm) was 0.37 compared to 0.17 for the observed EQ-5D-3L utility scores, suggesting the mapped EQ-5D-3L is more responsive to change.

**Conclusions:** We have developed a simple, responsive, and robust mapping algorithm to enable EQ-5D-3L health utilities to be generated from 10 items of the C19-YRSm. This mapping algorithm will facilitate economic evaluations of interventions, treatment, and management of people with LC, as well as further helping to describe and characterise patients with LC irrespective of any treatment and interventions.

## Background

Long Covid (LC) is a clinical syndrome of persistent symptoms beyond four weeks after a confirmed or possible COVID-19 infection.^1^ The term was coined by patients and encompasses the National Institute for Health and Care Excellence (NICE) defined terms, ‘ongoing symptomatic COVID-19’ (persistent symptoms 4-12 weeks after the infection) and ‘Post-COVID Syndrome (PCS)’ (symptoms persisting beyond 12 weeks after infection).^2,3^ As per the last official survey, there are an estimated 2 million cases of LC in the UK alone and an estimated 200 million cases worldwide.^4^ LC is a multisystem syndrome with more than 200 symptoms reported across 10 organ systems with common symptoms being fatigue, pain, breathlessness, brain fog (cognitive problems), sleep problems, anxiety, and depression.^5^ It is a fluctuating condition with a protracted course in some individuals causing significant distress and disability to the individual.^6^ LC has detrimental effects on health-related quality of life (HRQoL) and activities of daily living, including individual’s work productivity and potentially their caregivers.^7,8^

A number of condition-specific patient-reported outcome measures (PROMs) have been developed to capture the multi-symptom nature of the condition and its impact on HRQoL.^9–12^ The COVID-19 Yorkshire Rehabilitation Scale (C19-YRS) was one of the first condition-specific HRQoL measures for LC reported in the literature, and provides a symptom severity score, functional disability score, other symptoms score, and overall health score.^13–15^ Following a psychometric analysis of the C19-YRS, a modified version of the measure is now in use (C19-YRSm).^9^ The validity, reliability and responsiveness of the C19-YRSm has been demonstrated to be satisfactory in recent studies with excellent internal consistency.^16–20^

Although PROMs such as the C19-YRSm provide measures of HRQoL in LC, health technology assessment (HTA) agencies that focus on the evaluation of interventions, health service planners require preference-based HRQoL measures that generate health utilities^21^ for the purposes of cost-utility analyses. Health utilities describe HRQoL on a metric indexed at 0, representing death, and 1, representing perfect or full health, whereas utilities < 0 reflect states deemed worse than dead. One of the most commonly employed preference-based HRQoL measures is the EQ-5D.^22,23^ The EQ-5D generates generic health utilities applicable to general and clinical populations. This enables the comparison of interventions targeting different diseases in terms of their relative impact on HRQoL, measured using the EQ-5D, and survival benefits. Although the EQ-5D has been used in LC^24,25^ there may be instances necessitating health utilities, such as for health economic evaluations, where data have been collected with condition-specific PROMs, but not the EQ-5D. Furthermore, there is some recent evidence that has suggested that the EQ-5D, may not be as sensitive or responsive to change in health status among LC patients as the condition-specific C19-YRSm.^20^ An expedient is therefore needed to translate scores from the LC-specific C19-YRSm to the EQ-5D to derive health utilities. Mapping or cross-walking is one such approach enabling^26^ health utilities to be obtained from condition-specific, non-preference-based HRQoL measures.^27, 28^

There is, consequently, a need to evaluate the HRQoL of LC patients derived from generic and condition-specific measures, and explore the mechanism by which HRQoL outcomes derived from condition-specific measures may be transformed into health utilities for health economic evaluations. The aim of this study was therefore to compare EQ-5D with C19-YRSm and to develop a mapping algorithm for the C19-YRSm onto the EQ-5D to enable utility values to be generated from responses to the C19-YRSm.

## Methods

The data for this study were collected from the LOng COvid Multidisciplinary consortium Optimising Treatments and servIces acrOss the NHS (LOCOMOTION) study.^29^ LOCOMOTION is a prospective mixed-methods study involving 10 Long Covid services across the United Kingdom. Ethics approval for the LOCOMOTION study was obtained from the Bradford and Leeds Research Ethics Committee on behalf of Health Research Authority and Health and Care Research Wales (reference: 21/YH/0276).

Participants with a diagnosis of LC receiving treatment and management of the condition from one of the 10 participating LC services were recruited and registered on a digital PROM (DPROM) platform developed by the digital health company ELAROS 24/7 Ltd (ELAROS) and the University of Leeds.^30^ Although different services offered a variety of different PROMs via the platform, the C19-YRSm and EQ-5D-5L were the minimum number of PROMs each participant had to complete at registration and every 3 months thereafter. Participants received reminders via the platform portal to complete the PROMs.

### Instruments

#### Covid-19 Yorkshire Rehabilitation Scale – Modified (C19-YRSm)

The C19-YRSm is a 17-item instrument^9^ designed to capture the key symptoms of LC and its impact on activities of daily living and overall health. The items comprise four subscales: Symptom Severity (SS, 10 scored items), Functional Disability (FD, 5 items), Overall Health (OH, a single item), and Other Symptoms (OS). Previous research has demonstrated the C19-YRSm to be a well-validated PROM with robust psychometric properties.^16,20^

The items in the SS subscale comprise the following domains: breathlessness (4 items), cough/throat sensitivity/voice change (2 items), fatigue (one item), smell / taste (2 items), pain / discomfort (five items), cognition (three items), palpitations / dizziness (two items), post-exertional malaise (one item), anxiety / mood (five items), and sleep (one item). FD consists of 5 single items: communication, walking / moving around, personal care, other activities of daily living, and social role. Although the SS subscale contains 26 items, these are scored as 10 ‘domain’ items, where the highest value within each of the domains is used (e.g., breathlessness, pain / discomfort)

Responses on the SS and FD subscales are rated on a 0 (no symptom or dysfunction) to 3 (severe life-disturbing symptom or dysfunction) Likert scale. Higher scores on both these subscales indicate worse symptomatology and poorer functioning. Responses on the OH subscale are scored on a 0-10 Likert scale (0 being “worst health” and 10 being “best health”) with higher scores indicating better health. OS over the last 7 days are also captured from a list of 26 additional symptoms.^9^

#### The EuroQol 5D-5L (EQ-5D-5L)

The EuroQol EQ-5D-5L is a preference-based HRQoL instrument with five dimensions: Mobility, Usual Activities, Selfcare, Pain / Discomfort, and Anxiety / Depression.^22^ It has five response categories for each dimension ranging from 1 (no problems) to 5 (severe problems). Responses to each dimension are collated into a profile score, which is converted into a health utility or index score using a country-specific algorithm (tariff or value set). Utilities reflect societal preferences for health states and are measured on a metric indexed at 0 (dead) to 1 (perfect health). Utility values less than 0, indicating states worse than dead, are also captured. The EQ-5D-5L also comprises a visual analogue scale (VAS) measuring self-reported current health on a scale from 0 (“worst health”) to 100 (“best health”). The EQ-VAS was not used in the mapping analysis.

The EQ-5D-5L scores were converted into EQ-5D-3L utilities^23^ using the crosswalk (CW) algorithm to derive UK utility values.^31^ The UK utility values were derived using the approach recommended by NICE, which currently consists of applying a validated mapping function onto the UK EQ-5D-3L tariff set that has been developed by the NICE Decision Support Unit.^32^

### Data

Data for the current research were collated from the LOCOMOTION study. Two datasets were derived for the analysis. For the primary analysis, i.e., derivation of the mapping algorithm, data were extracted for patients who had completed the C19-YRSm and EQ-5D-5L only once (at the same assessment point, i.e., on the same day). This sample was then randomly split into two further, equally numbered, subsets to create estimation and validation subsets for the mapping algorithm.

A second sample was generated from patients with multiple, contemporaneous assessments on both the C19-YRSm and EQ-5D-5L. This sample was used to evaluate the responsiveness, or sensitivity to change, of the mapped health utility values derived from the C19-YRSm relative to those of the actual EQ-5D-5L utilities (see below).

### Statistical analysis

Basic clinical and sociodemographic details were extracted from the primary dataset (participants with a single set of assessments) and summarised in frequency counts and percentages. Scores on the C19-YRSm domains (SS, FD, and OH) and EQ-5D-3L were summarised for the first assessment (“baseline”) using means, standard deviations (SD) and 95% confidence intervals (CIs).

The development and reporting of the mapping algorithm was undertaken in accordance with the MAPS (MApping onto Preference-based measures reporting Standards) statement.^33^ Spearman rank correlation was used to evaluate the level of association between the individual C19-YRSm items and the EQ-5D-3L index (utility) scores. C19-YRSm items with moderate to high levels of association with the EQ-5D-3L utility scores (r > 0.5) were included in the mapping models (see below).

In addition to this, univariate linear (ordinary least squares, OLS) regression models (using the training data subset) were applied to potential predictors from the sociodemographic and clinical variables: age, sex, ethnicity, number of pre-COVID comorbid medical conditions, as well as OH from the C19-YRSm. OS was omitted from this analysis as this optional subscale – which lists a total of 26 symptoms potentially affecting individuals with LC – is not always implemented or utilised in clinical practice. The statistical significance for the included variables was evaluated at p<0.10 for inclusion in the regression models.

Two OLS regression models were evaluated: 1) the full model comprising all C19-YRSm items, OH, and the sociodemographic and clinical variables; 2) the parsimonious model based on items from the C19-YRSm (r > 0.5) and OH.

Model fit was evaluated with the following: R^2^ and adjusted R^2^, Akaike Information Criteria (AIC) and log-likelihood statistics. The log likelihood ratio test was used to determine relative fit between the two models. The test subset was then utilised to assess the final mapping algorithm. Model fit of the predicted EQ-5D-3L mapped utility values was evaluated using R^2^, mean absolute error (MAE), and the root mean squared error (RMSE) values. Bootstrapping (10000 replications) was applied to evaluate the RMSE.

The mean difference between predicted and actual EQ-5D-3L health utilities was derived for each of the following range of scores of the EQ-5D-3L utilities to determine how well the predicted values corresponded to actual values across the range of scores: less than −0.2; −0.2 to < 0; ≥ 0 to < 0.4; ≥ 0.4 to < 0.6; ≥ 0.6 to < 0.8; and ≥ 0.8.

The final mapping algorithm was further validated by Bland-Altman plots^34^ for the actual and predicted EQ-5D-3L health utilities (by plotting the difference between the two forms of health utilities against the mean health utilities). The 95% limits of agreement between the two utility scores were derived (±2SD) as well as the percentage of difference scores falling outside these limits.

Finally, the responsiveness of the mapped EQ-5D-3L health utilities was assessed relative to the actual values using the second dataset. Data were extracted from participants with a first assessment and follow-up assessment at 90 days ± 10 days on both instruments. The mapping algorithm was applied to the C19-YRSm variables to derive mapped EQ-5D-3L utility values. Responsiveness was determined through the standardised response mean (SRM) by dividing the change from baseline by the SD of the change. The SRM was used to determine the relative responsiveness of the mapped to actual EQ-5D-3L utilities with higher SRM values corresponding to greater levels of responsiveness.

## Results

### Participants

A total of 1434 participants had completed both the C19-YRSm and EQ-5D-3L at a single time point (“first assessment”). The majority of participants were of white ethnicity (87%) and female (68%); the mean age for the sample was 46 years (standard deviation, SD: 14.2 years) (Table 1), and most participants (75%) had no comorbidities prior to LC. The average length of time between the first COVID-19 infection and the first assessment was approximately 430 days (SD: 281 days). A small number of participants had been hospitalised (12%) and admitted to intensive care (3%) as a result of COVID-19 infection. Just over a fifth (22%) had been discharged from the LC services. The mean domain scores on the C19-YRSm and EQ-5D-3L indicated a high level of symptom burden and negative impact on HRQoL resulting from LC.

**Table 1.**
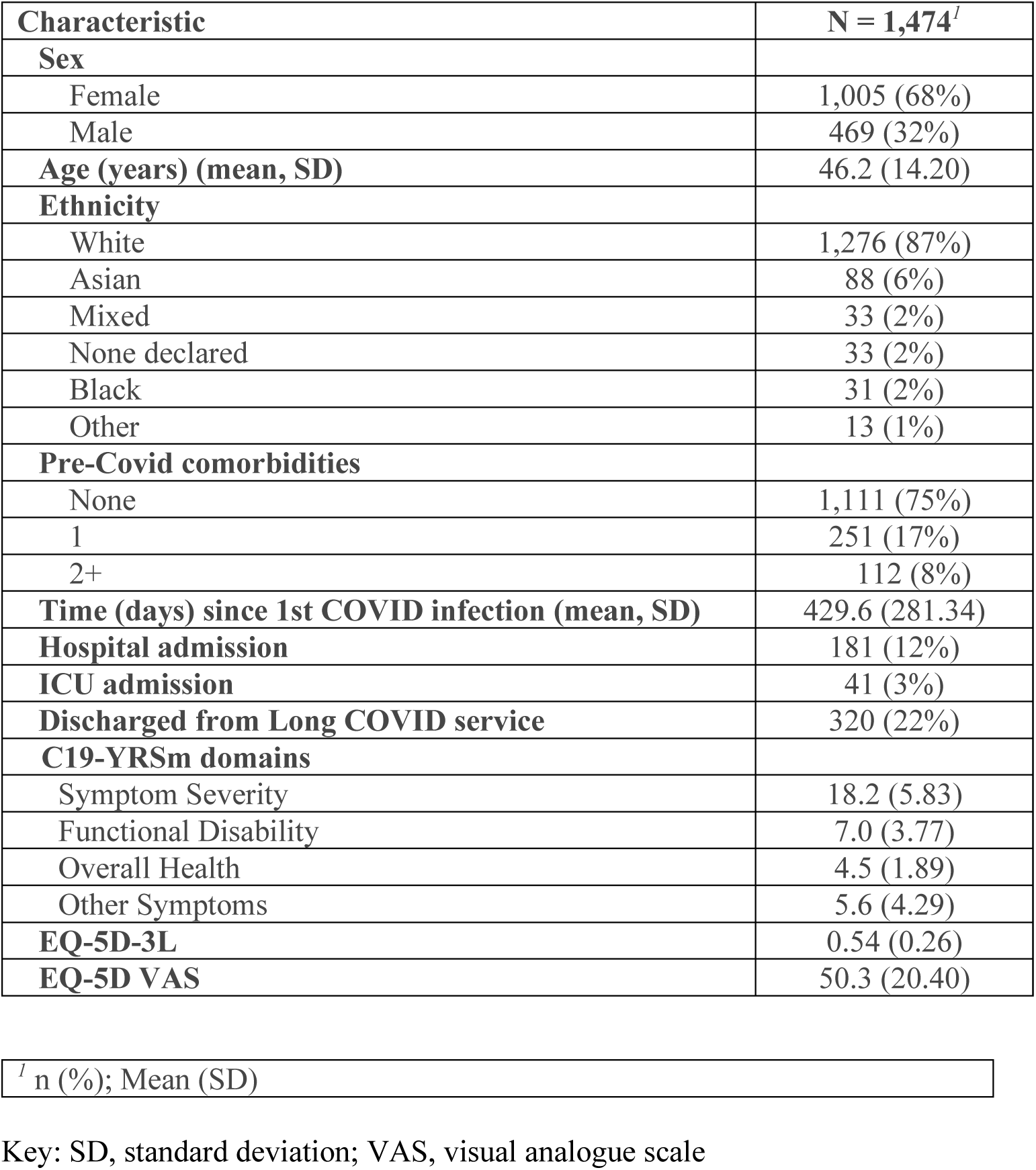
Clinical and sociodemographic details.

| **Characteristic** | **N = 1,474<sup>I</sup>** |
| --- | --- |
| **Sex** |  |
| Female | 1,005 (68%) |
| Male | 469 (32%) |
| **Age (years) (mean, SD)** | 46.2 (14.20) |
| **Ethnicity** |  |
| White | 1,276 (87%) |
| Asian | 88 (6%) |
| Mixed | 33 (2%) |
| None declared | 33 (2%) |
| Black | 31 (2%) |
| Other | 13 (1%) |
| **Pre-Covid comorbidities** |  |
| None | 1,111 (75%) |
| 1 | 251 (17%) |
| 2+ | 112 (8%) |
| **Time (days) since 1st COVID infection (mean, SD)** | 429.6 (281.34) |
| **Hospital admission** | 181 (12%) |
| **ICU admission** | 41 (3%) |
| **Discharged from Long COVID service** | 320 (22%) |
| **C19-YRSm domains** |  |
| Symptom Severity | 18.2 (5.83) |
| Functional Disability | 7.0 (3.77) |
| Overall Health | 4.5 (1.89) |
| Other Symptoms | 5.6 (4.29) |
| **EQ-5D-3L** | 0.54 (0.26) |
| **EQ-5D VAS** | 50.3 (20.40) |

<sup>I</sup> n (%); Mean (SD)
Key: SD, standard deviation; VAS, visual analogue scale

### Mapping-Potential Predictors

The results of the univariate linear regression analyses revealed the C19-YRSm OH (beta=0.07, standard error (se) 0.004, p<0.0001) subscale was a statistically significant predictor for the EQ-5D-3L utility values. Similarly, statistically significant results were also shown for Sex (beta=0.023, se= 0.02, p=0.01) and pre-LC comorbidities (beta=-0.069, se=0.015, p=0.0002). Neither Ethnicity nor Age were statistically significant (p=0.31; p=0.44, respectively). Nevertheless, as potentially important covariates, these were included in the full model alongside Sex, pre-LC comorbidities, and OH.

### Mapping Algorithm

The training set for the mapping algorithm comprised 737 participants. The distribution of the EQ-5D-3L health utilities is shown in Figure 1. The mean EQ-5D-3L health utility value was 0.534 (SD: 0.266, minimum: −0.346, maximum: 1; 95%CI: 0.52; 0.56). The interquartile range (25^th^ to 75^th^ centile) was 0.378 to 0.728.

**Figure 1.**
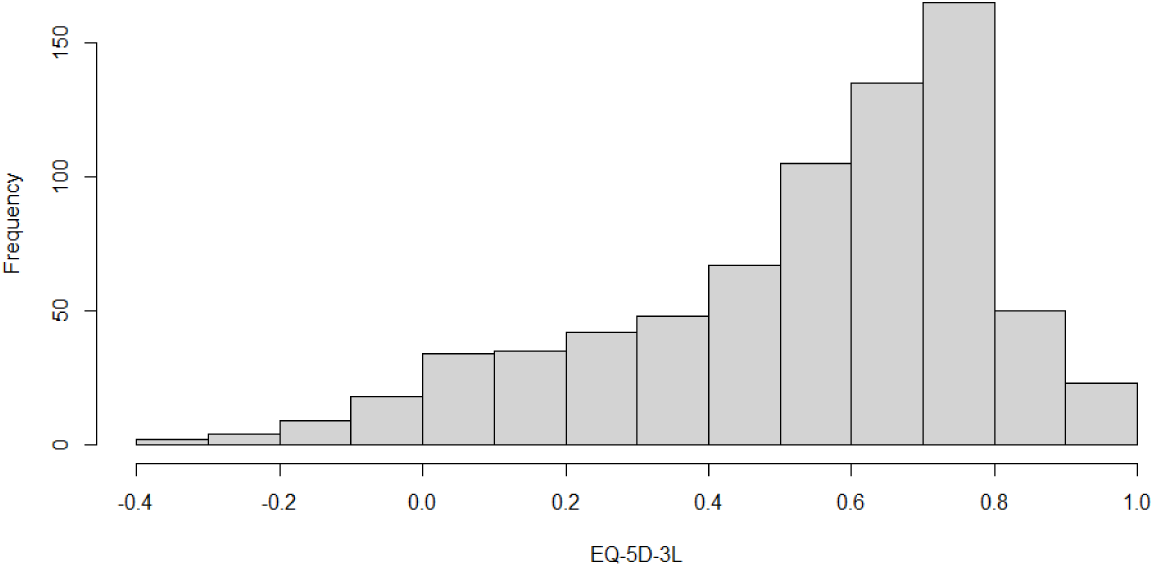
Distribution of the EQ-5D-5L Index Scores.

The results of the correlation analysis Supplementary Table 1 showed moderate-to-high levels of association between the FD items and EQ-5D-3L utility scores: “Walking / Moving around” (item Q12, r −0.63), “Personal care” (Q13, r −0.66), “Other Activities of Daily Living” (Q14, r −0.55) and “Social role” (Q15, r −0.58). Similarly, “Muscle pain” (Q5C), “Feeling depressed” (Q9B), and “Breathlessness (on dressing yourself)” (Q1C) also showed good levels of association with the EQ-5D-3L utility scores. “Joint pain” (Q5B) and “Feeling anxious” (Q9A) fell just short of the 0.5 criterion, however, but were included in the model due to the conceptual overlap with the EQ-5D-3L dimensions (“Pain / discomfort” and “Anxiety / Depression”, respectively).

The results of the full and parsimonious models are shown in Supplementary Table 2 and Table 2, respectively. For the full model, R^2^ was 0.71 and the adjusted R^2^ 0.67 for this model; for the parsimonious model these were R^2^ 0.68 and adjusted R^2^ 0.67, respectively. The log-likelihood ratio test showed no statistically significant differences in terms of goodness of fit between the two models (X^2^ 82.2, p=0.32). The parsimonious model was therefore selected as the most appropriate mapping model. The mapping algorithm has been provided in Appendix 1.

**Table 2.**
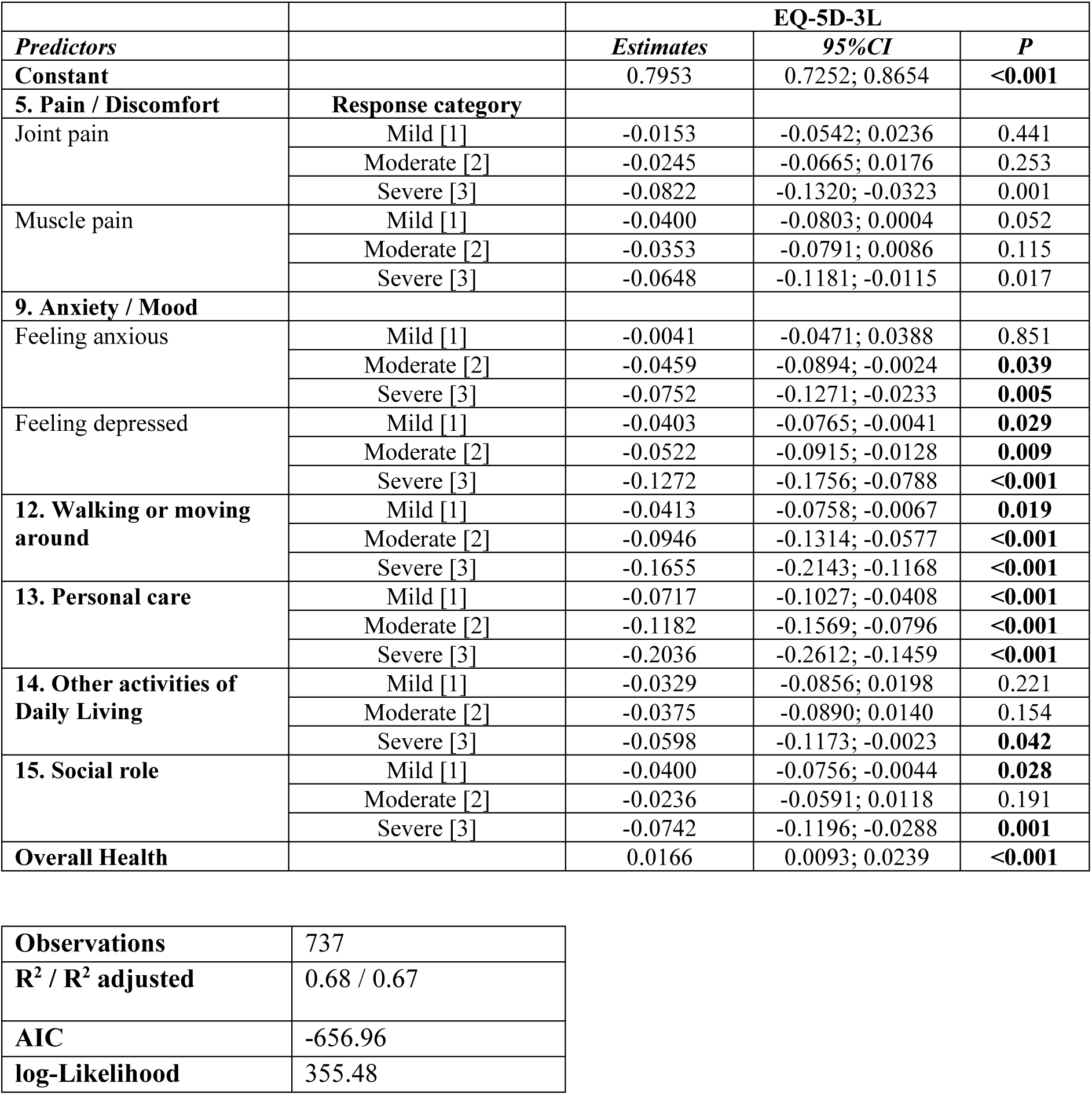
Regression coefficients for the Final (Parsimonious) Model.

|  |  | **EQ-5D-3L** |  |  |
| --- | --- | --- | --- | --- |
| *Predictors* |  | *Estimates* | *95%CI* | *P* |
| **Constant** |  | 0.7953 | 0.7252; 0.8654 | **<0.001** |
| **5. Pain / Discomfort** | **Response category** |  |  |  |
| Joint pain | Mild [1] | -0.0153 | -0.0542; 0.0236 | 0.441 |
|  | Moderate [2] | -0.0245 | -0.0665; 0.0176 | 0.253 |
|  | Severe [3] | -0.0822 | -0.1320; -0.0323 | 0.001 |
| Muscle pain | Mild [1] | -0.0400 | -0.0803; 0.0004 | 0.052 |
|  | Moderate [2] | -0.0353 | -0.0791; 0.0086 | 0.115 |
|  | Severe [3] | -0.0648 | -0.1181; -0.0115 | 0.017 |
| **9. Anxiety / Mood** |  |  |  |  |
| Feeling anxious | Mild [1] | -0.0041 | -0.0471; 0.0388 | 0.851 |
|  | Moderate [2] | -0.0459 | -0.0894; -0.0024 | **0.039** |
|  | Severe [3] | -0.0752 | -0.1271; -0.0233 | **0.005** |
| Feeling depressed | Mild [1] | -0.0403 | -0.0765; -0.0041 | **0.029** |
|  | Moderate [2] | -0.0522 | -0.0915; -0.0128 | **0.009** |
|  | Severe [3] | -0.1272 | -0.1756; -0.0788 | **<0.001** |
| **12. Walking or moving around** | Mild [1] | -0.0413 | -0.0758; -0.0067 | **0.019** |
|  | Moderate [2] | -0.0946 | -0.1314; -0.0577 | **<0.001** |
|  | Severe [3] | -0.1655 | -0.2143; -0.1168 | **<0.001** |
| **13. Personal care** | Mild [1] | -0.0717 | -0.1027; -0.0408 | **<0.001** |
|  | Moderate [2] | -0.1182 | -0.1569; -0.0796 | **<0.001** |
|  | Severe [3] | -0.2036 | -0.2612; -0.1459 | **<0.001** |
| **14. Other activities of Daily Living** | Mild [1] | -0.0329 | -0.0856; 0.0198 | 0.221 |
|  | Moderate [2] | -0.0375 | -0.0890; 0.0140 | 0.154 |
|  | Severe [3] | -0.0598 | -0.1173; -0.0023 | **0.042** |
| **15. Social role** | Mild [1] | -0.0400 | -0.0756; -0.0044 | **0.028** |
|  | Moderate [2] | -0.0236 | -0.0591; 0.0118 | 0.191 |
|  | Severe [3] | -0.0742 | -0.1196; -0.0288 | **0.001** |
| **Overall Health** |  | 0.0166 | 0.0093; 0.0239 | **<0.001** |

|  |  |
| --- | --- |
| **Observations** | 737 |
| **R<sup>2</sup> / R<sup>2</sup> adjusted** | 0.68 / 0.67 |
| **AIC** | -656.96 |
| **log-Likelihood** | 355.48 |

### Predicted EQ-5D-3L

The test set for the mapping algorithm comprised 734 participants. The mean EQ-5D-3L health utility value was 0.54 (SD: 0.264, minimum: −0.346, maximum: 1; 95%CI: 0.52, 0.56). The mean predicted EQ-5D-3L health utility was 0.533 (SD: 0.222, minimum: −0.0571, maximum: 0.945; 95%CI: 0.52, 0.55). The R^2^ was 0.68 and RMSE 0.15 (Figure 2).

**Figure 2.**
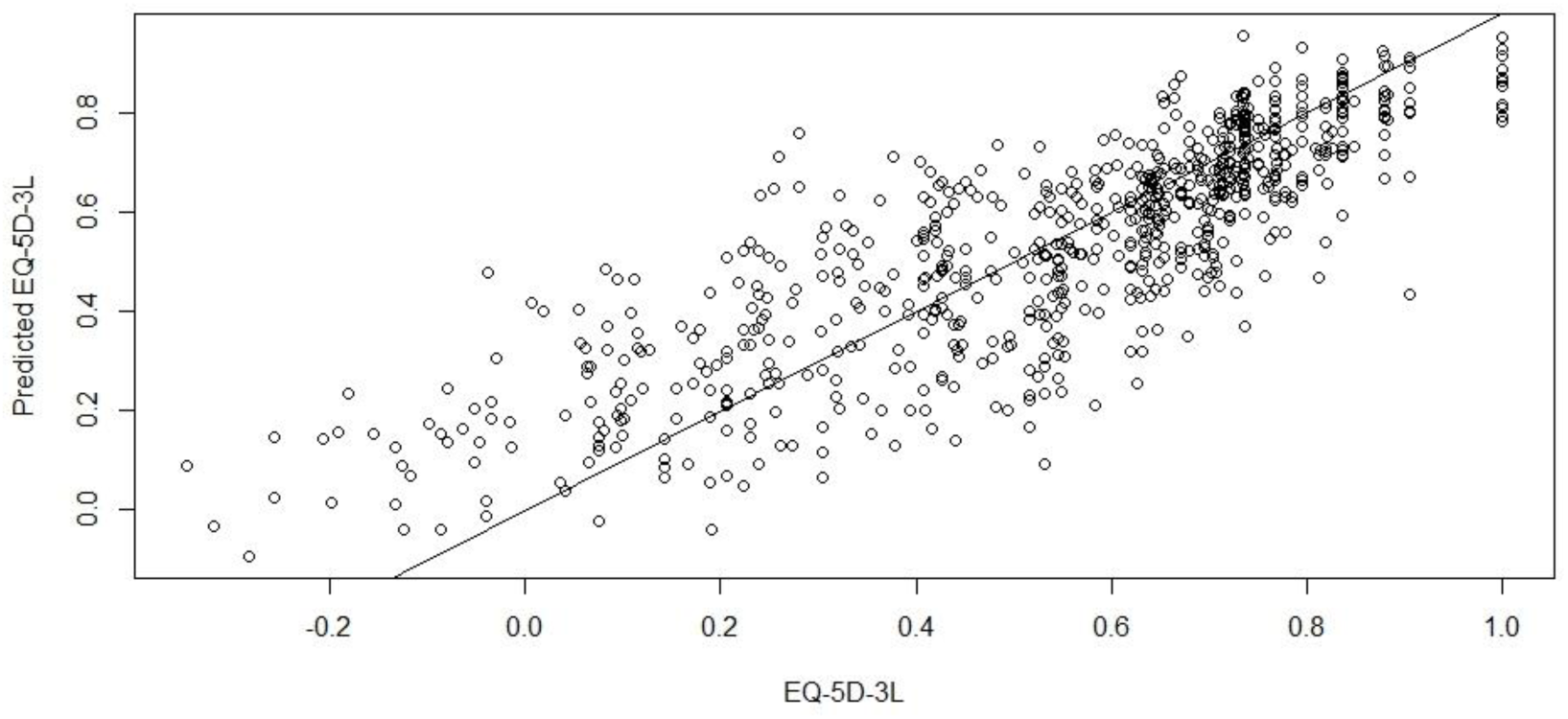
Actual versus Predicted EQ-5D-3L Index scores.

Table 3 shows the mean difference between the actual and predicted EQ-5D-3L health utilities across the range of (observed) EQ-5D-3L utility values. The mean difference between the two sets of scores was ≤ 0.10 for the range from 0 to 1 suggesting a good degree of targeting for positive values of the EQ-5D-3L. The mean differences for values less than 0 were greater, suggesting poorer targeting, which may, however, have been impacted by the smaller number of participants with poor health utilities. The Bland-Altman plot is shown in Figure 3. The mean difference between scores was 0.007 (SD: 0.15; 95%CI: −0.004, 0.012) and the limits of agreement were −0.293 and 0.307.

**Figure 3.**
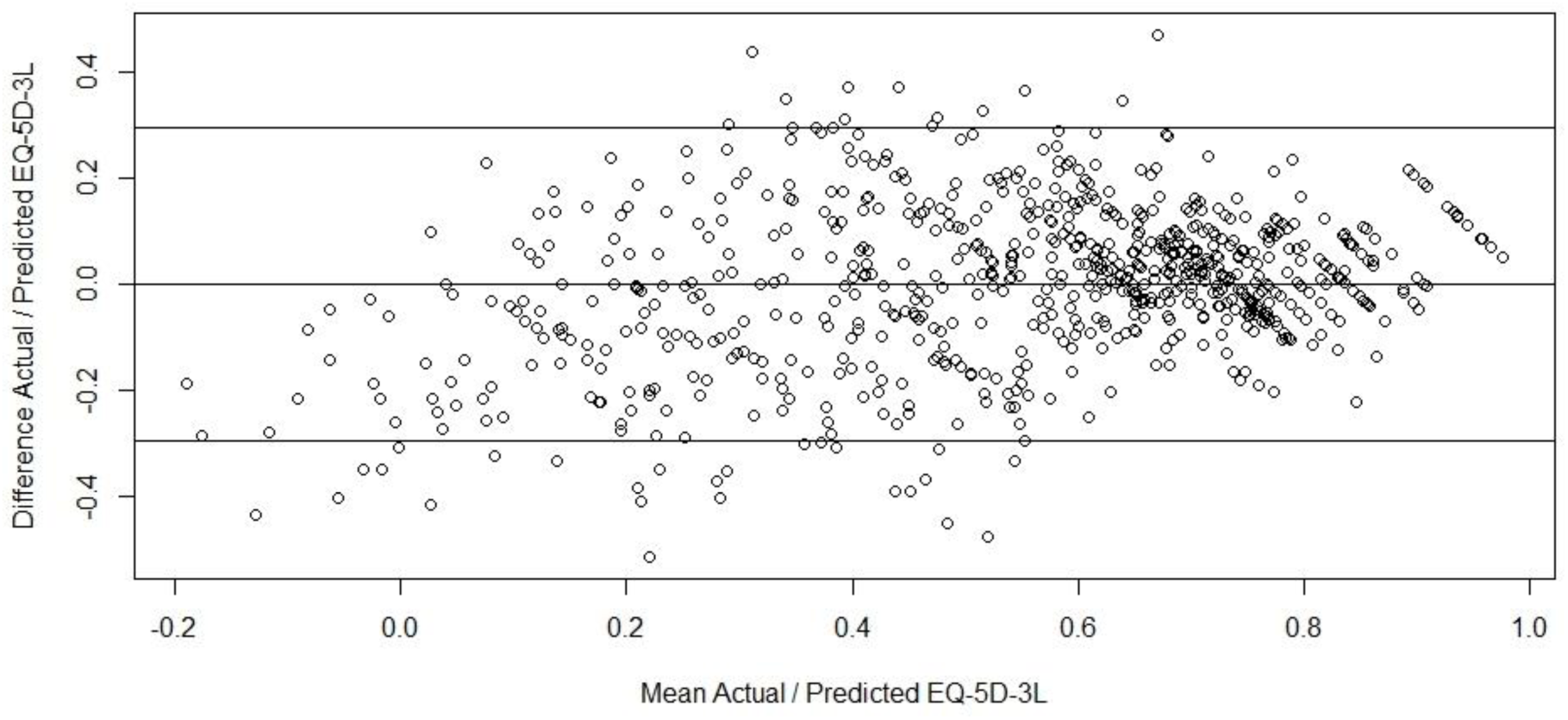
Bland-Altman Plot: Actual versus Predicted EQ-5D-3L.

**Table 3.** Mean difference between actual and predicted EQ-5D-3L by range of (actual) EQ-5D-3L.

| **Range EQ-5D-3L** | **<-0.2,**<br>**N = 6<sup>1</sup>** | **≥ 0.2 to**<br>**<0,**<br>**N = 26<sup>1</sup>** | **≥ 0 to**<br>**<0.20,**<br>**N = 59<sup>1</sup>** | **≥ 0.20 to**<br>**<0.40,**<br>**N = 94<sup>1</sup>** | **≥ 0.40 to**<br>**<0.60,**<br>**N = 164<sup>1</sup>** | **≥ 0.60 to**<br>**<0.80,**<br>**N = 305<sup>1</sup>** | **≥ 0.80,**<br>**N = 83<sup>1</sup>** |
| --- | --- | --- | --- | --- | --- | --- | --- |
| **Mean difference** | -0.34<br>(0.09) | -0.21<br>(0.13) | -0.13<br>(0.14) | -0.08<br>(0.17) | 0.02<br>(0.14) | 0.05<br>(0.10) | 0.09<br>(0.10) |

<sup>1</sup> Mean (SD)

Finally, there were 85 participants who had completed both instruments at first assessment and 90 days later (± 10 days). The mapping algorithm (Appendix 1) was applied to the C19-YRSm variables. Table 4 shows the mean values for the mapped and observed EQ-5D-3L utility scores. It may be seen than at first assessment the mapped EQ-5D-3L utility values are equal to the observed EQ-5D-3L utility values. At follow-up, both sets demonstrated an improvement in mean health utilities over time with the mapped values showing slightly greater improvement (0.02). The SRM was 0.37 for the mapped EQ-5D-3L health utilities compared to 0.17 for the actual EQ-5D-3L utility scores, suggesting the mapped EQ-5D-3L (based on the C19-YRSm) is more responsive to change.

**Table 4.** Mean EQ-5D-3L indices (mapped and actual)

| **Mean EQ-5D-3L (SD)**<br>**[95%CI] N = 85<sup>1</sup>** | **Actual** | **Mapped** |
| --- | --- | --- |
| First assessment | 0.55 (0.22) [0.50, 0.60] | 0.55 (0.21) [0.50, 0.59] |
| Follow-up (90 days ±<br>10 days) | 0.58 (0.23) [0.53, 0.63] | 0.60 (0.19) [0.56, 0.64] |

<sup>1</sup> Mean (SD); 95% confidence interval

## Discussion

The aim of this study was to derive a mapping algorithm to generate preference-based EQ-5D-3L health utilities from the condition-specific C19-YRSm in people with LC. The mapping algorithm demonstrated very good face validity in terms of the final items and domains from the C19-YRSm selected with a high degree of correspondence with the single-item dimensions on the EQ-5D-3L. The items from the C19-YRSm in the final model, such as walking or moving around (item 12A), personal care (13A), social role (15A), as well as anxiety, depression (9A, 9B), and (joint and muscle) pain (5B and C), strongly overlapped with the corresponding dimension items in the EQ-5D. OH and OS were two other C19-YRSm items in the mapping algorithm capturing the more general HRQoL aspects of people experiencing LC.

The final mapping algorithm model showed good overall fit on the criterion indices (R^2^ and RMSE). There was furthermore a high degree of agreement (little bias) between the predicted and actual EQ-5D-3L health utilities with approximately 5% of the differences between these estimates falling outside 2 standard deviations. The results of the final analysis revealed that the SRM of mapped utility values were greater than those for the observed EQ-5D-3L utility values, suggesting that the mapped measures were more responsive to change. Although the latter should perhaps be interpreted with a degree of caution given the relatively small sample size, it nevertheless underlines the fact that condition-specific PROMs may be more sensitive and responsive to symptom and functional changes being experienced by people with LC than generic PROMs.

Some potential limitations should be highlighted, including the fact that both the estimation and validation data were drawn from the same sample with no external validation set. This may have introduced a degree of bias in the parameter estimates. A possible concomitant of this was that the targeting of the mapped EQ-5D health utilities was less optimal at the lower end of the observed EQ-5D utility range, particularly for utility values less than −0.20. However, this may also reflect the distribution of both the C19-YRSm scores, and particularly the EQ-5D-3L utility values with fewer people with LC experiencing health states as severe as “worse than dead” (i.e., less than 0), as only 5% (N=33) of the sample had observed EQ-5D-3L utility values < 0. This is an area to be explored further in future research.

The use of a single regression (OLS) may be considered a possible shortcoming, however it is unlikely that model fit would have been improved significantly beyond the high levels demonstrated in this study through the application of other regression models.^26^ Therefore, taken in the round, the large sample size, concordance between items from the C19-YRSm in the mapping algorithm and the EQ-5D single-item dimensions, the high level of variance explained (“fit”) by the mapping model, as well as the degree of agreement between utility estimates all underpin the robustness of the results.

These results have important implications for the use of C19-YRsm in clinical practice and service evaluation. LC is by its nature is a varying condition with daily fluctuations in both symptom and symptom severity.^35^ The results of this study emphasise the need for COVID-specific measures when evaluating HRQoL, particularly in the context of determining the cost-effectiveness of interventions and healthcare services aimed at LC.

Despite the evident correspondence between the EQ-5D-5L single-item dimensions and the C19-YRSm, the generic instrument may not fully capture the HRQoL of people experiencing LC, nor specifically be responsive to the variable nature of the condition. LC has been shown to be associated with a loss of independence^36^, which in turn has a significant negative impact, on work ability and productivity.^7,8^

Randomised controlled trials evaluating interventions aimed at supporting and improving the ability of people with LC to return to work^37^ require specific instruments, such as the C19-YRSm, that are able to measure the benefits of these interventions and are both relevant and specific to patients by measuring what is important to them. Yet, at the same time, health policy, particularly in jurisdictions where health care is funded from the public purse, requires evidence of the cost-effectiveness of interventions^38,39^ supported by an instrument sensitive and responsive to the fluctuating nature of LC. Clearly, the ability to derive health utilities including the EQ-5D utility scores from the C19-YRSm could play an important role in this.

## Conclusion

We have developed a simple, responsive, and robust mapping algorithm to enable EQ-5D-3L health utilities to be generated from 10 items of the C19-YRSm. This mapping algorithm will facilitate the health economic evaluation of interventions, treatment, and management of people with LC.

## Data Availability

All data produced in the present work are contained in the manuscript.

## Appendix 1. Mapping algorithm C19-YRSm

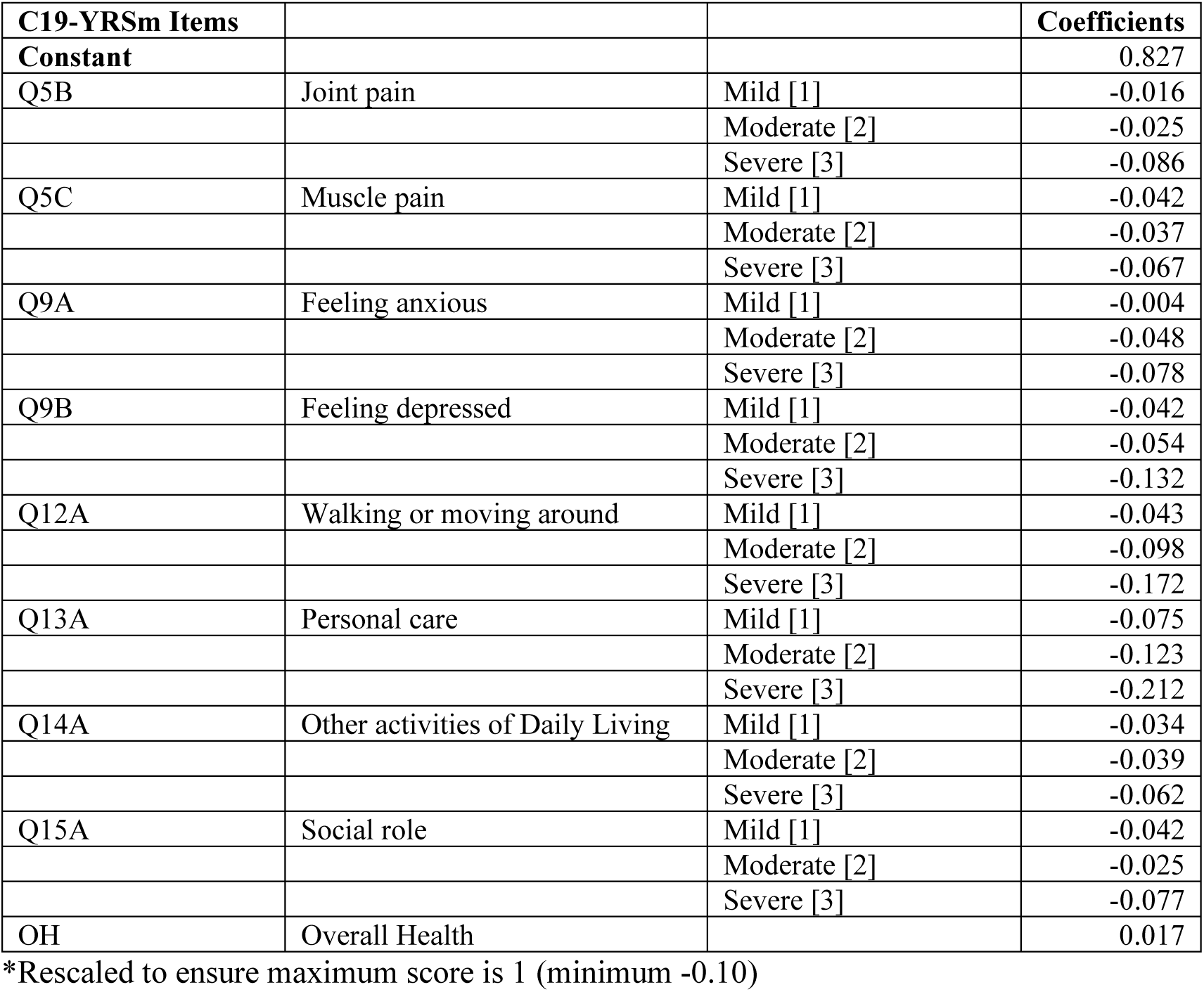

| C19-YRSm Items |  |  | Coefficients |
| --- | --- | --- | --- |
| Constant |  |  | 0.827 |
| Q5B | Joint pain | Mild [1] | -0.016 |
|  |  | Moderate [2] | -0.025 |
|  |  | Severe [3] | -0.086 |
| Q5C | Muscle pain | Mild [1] | -0.042 |
|  |  | Moderate [2] | -0.037 |
|  |  | Severe [3] | -0.067 |
| Q9A | Feeling anxious | Mild [1] | -0.004 |
|  |  | Moderate [2] | -0.048 |
|  |  | Severe [3] | -0.078 |
| Q9B | Feeling depressed | Mild [1] | -0.042 |
|  |  | Moderate [2] | -0.054 |
|  |  | Severe [3] | -0.132 |
| Q12A | Walking or moving around | Mild [1] | -0.043 |
|  |  | Moderate [2] | -0.098 |
|  |  | Severe [3] | -0.172 |
| Q13A | Personal care | Mild [1] | -0.075 |
|  |  | Moderate [2] | -0.123 |
|  |  | Severe [3] | -0.212 |
| Q14A | Other activities of Daily Living | Mild [1] | -0.034 |
|  |  | Moderate [2] | -0.039 |
|  |  | Severe [3] | -0.062 |
| Q15A | Social role | Mild [1] | -0.042 |
|  |  | Moderate [2] | -0.025 |
|  |  | Severe [3] | -0.077 |
| OH | Overall Health |  | 0.017 |

\*Rescaled to ensure maximum score is 1 (minimum -0.10)

### Mapping algorithm

Utility = 0.779+(Q5B+Q5C+Q9A+Q9B+Q12A+Q13A+Q14A+Q15A+(OH*0.017))

For instance, a patient with moderate (score 2) joint and muscle pain (Q5A, Q5B), mild anxiety and depression (score 1) (Q9A and Q9B), severe issues (score 3) with walking (Q12A), personal care (Q13A), activities of daily living (Q14A) and social role (Q15A), with an Overall Health score of 5 and 6 Other Symptoms:

Utility = 0.827+(−0.025)+(−0.037)+(−0.004)+(−0.042)+(−0.172)+(−0.212)+(−0.062)+(−0.077)+(5*0.017)

Utility = 0.282

## Appendix 2

The C19-YRSm is available at: https://c19-yrs.com/wp-content/uploads/2024/07/Modified-C19YRS-Self-Report.pdf

## List of abbreviations

AIC: Akaike Information Criteria

C19-YRSm: Covid 19 Yorkshire Rehabilitation Screening Tool (modified)

CI: Confidence Interval

EQ-5D-3L: EuroQol 5-Dimension 3-Level

EQ-5D-5L: EuroQol 5-Dimension 5-Level

FD: Functional Disability (from the C19-YRSm)

HRQoL: Health-related Quality of Life

HTA: Health Technology Assessment

LC: Long COVID

MAPS: MApping onto Preference-based measures reporting Standards

NHS: National Health Service

NICE: National Institute for Health and Care Excellence

OH: Overall Health (from the C19-YRSm)

OLS: Ordinary Least Squares

OS: Other Symptoms (from the C19-YRSm)

PCS: Post-COVID Syndrome

PROM: Patient-reported Outcome Measure

RMSE: Root Mean Squared Error

SD: Standard Deviation

SE: Standard Error

SRM: Standardised Response Mean

SS: Symptom Severity (from the C19-YRSm)

VAS: Visual Analogue Scale

## Declarations

### Ethics approval and consent to participate

Ethics approval for the LOCOMOTION study was obtained from the Bradford and Leeds Research Ethics Committee on behalf of Health Research Authority and Health and Care Research Wales (reference: 21/YH/0276).

### Consent for publication

Not applicable.

### Availability of data and materials

All data generated or analysed during this study are included in this published article and its supplementary information files.

### Authors’ contributions

ABS analysed the data and drafted the manuscript; DCG reviewed the data analysis and co-drafted the manuscript; MS co-drafted the manuscript. All authors read and approved the final manuscript.

## Acknowledgements

The authors would like to thank all people with Long Covid and the health care professionals who participated in the study. We thank members of the digital health company ELAROS 24/7 Ltd who designed and developed the DPROM platform. We also thank the University of Leeds and EuroQol commercial teams for licensing the C19-YRSm (Yorkshire Rehabilitation Scale) and EQ-5D-5L scales respectively to ELAROS. The LOCOMOTION consortium: Nawar Diar Bakerly, Mauricio Barahona, Alexander Casson, Jonathan Clarke, Vasa Curcin, Helen Davies, Helen Dawes, Brendan Delaney, Carlos Echevarria, Sarah Elkin, Rachael Evans, Zaccheus Falope, Darren Greenwood Ben Glampson, Stephen Halpin, Mike Horton, Joseph Kwon, Simon de Lusignan, Gayathri Delanerolle, Erik Mayer, Harsha Master, Ruairidh Milne, Jacqui Morris, Amy Parkin, Anton Pick, Nick Preston, Amy Rebane, Emma Tucker, Ana Belen Espinosa Gonzalez, Sareeta Baley, Annette Rolls, Emily Bullock, Megan Ball, Shehnaz Bashir, Mae Mansoubi, Joanne Elwin, Denys Prociuk, Iram Qureshi, Samantha Jones.

## Authors’ information (optional)

–

---

**Supplementary Table 1.**
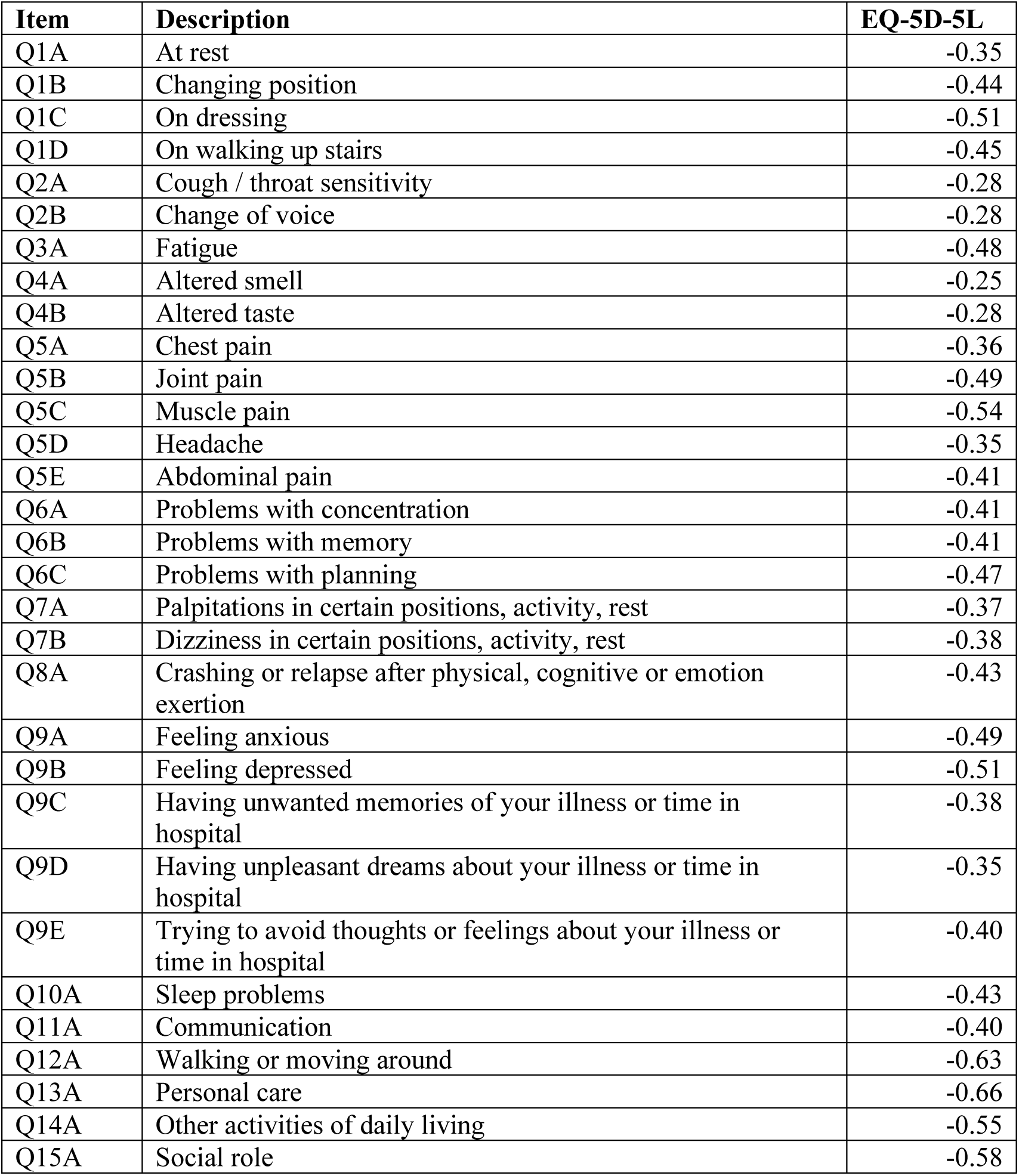
Correlation analysis: C19-YRSm items and EQ-5D-5L.

| **Item** | **Description** | **EQ-5D-5L** |
| --- | --- | --- |
| Q1A | At rest | -0.35 |
| Q1B | Changing position | -0.44 |
| Q1C | On dressing | -0.51 |
| Q1D | On walking up stairs | -0.45 |
| Q2A | Cough / throat sensitivity | -0.28 |
| Q2B | Change of voice | -0.28 |
| Q3A | Fatigue | -0.48 |
| Q4A | Altered smell | -0.25 |
| Q4B | Altered taste | -0.28 |
| Q5A | Chest pain | -0.36 |
| Q5B | Joint pain | -0.49 |
| Q5C | Muscle pain | -0.54 |
| Q5D | Headache | -0.35 |
| Q5E | Abdominal pain | -0.41 |
| Q6A | Problems with concentration | -0.41 |
| Q6B | Problems with memory | -0.41 |
| Q6C | Problems with planning | -0.47 |
| Q7A | Palpitations in certain positions, activity, rest | -0.37 |
| Q7B | Dizziness in certain positions, activity, rest | -0.38 |
| Q8A | Crashing or relapse after physical, cognitive or emotion exertion | -0.43 |
| Q9A | Feeling anxious | -0.49 |
| Q9B | Feeling depressed | -0.51 |
| Q9C | Having unwanted memories of your illness or time in hospital | -0.38 |
| Q9D | Having unpleasant dreams about your illness or time in hospital | -0.35 |
| Q9E | Trying to avoid thoughts or feelings about your illness or time in hospital | -0.40 |
| Q10A | Sleep problems | -0.43 |
| Q11A | Communication | -0.40 |
| Q12A | Walking or moving around | -0.63 |
| Q13A | Personal care | -0.66 |
| Q14A | Other activities of daily living | -0.55 |
| Q15A | Social role | -0.58 |

**Supplementary Table 2.** Regression coefficients for the Full Model.

| *Predictors* |  | **EQ-5D-3L** |  |  |
| --- | --- | --- | --- | --- |
|  |  | *Estimates* | *95%CI* | *P* |
|  | **Constant** | 0.7964 | 0.6720; 0.9209 | **<0.001** |
| **1. Breathlessness** | **Response category** |  |  |  |
| At rest | Mild [1] | -0.0180 | -0.0533; 0.0174 | 0.318 |
|  | Moderate [2] | -0.0158 | -0.0574; 0.0258 | 0.456 |
|  | Severe [3] | -0.0756 | -0.1512; -0.0000 | 0.050 |
| Changing position | Mild [1] | 0.0187 | -0.0218; 0.0592 | 0.366 |
|  | Moderate [2] | 0.0135 | -0.0377; 0.0647 | 0.605 |
|  | Severe [3] | 0.0795 | -0.0088; 0.1679 | 0.078 |
| On dressing | Mild [1] | -0.0193 | -0.0573; 0.0188 | 0.321 |
|  | Moderate [2] | -0.0271 | -0.0784; 0.0242 | 0.300 |
|  | Severe [3] | -0.0485 | -0.1242; 0.0272 | 0.209 |
| On walking up stairs | Mild [1] | 0.0500 | 0.0042; 0.0957 | 0.032 |
|  | Moderate [2] | 0.0287 | -0.0238; 0.0812 | 0.283 |
|  | Severe [3] | -0.0010 | -0.0632; 0.0612 | 0.974 |
| **2. Cough and/or throat sensitivity / voice change** |  |  |  |  |
| Cough / throat sensitivity | Mild [1] | 0.0105 | -0.0207; 0.0417 | 0.510 |
|  | Moderate [2] | -0.0045 | -0.0412; 0.0321 | 0.808 |
|  | Severe [3] | 0.0198 | -0.0358; 0.0755 | 0.484 |
| Change of voice | Mild [1] | 0.0107 | -0.0225; 0.0438 | 0.528 |
|  | Moderate [2] | 0.0267 | -0.0149; 0.0683 | 0.208 |
|  | Severe [3] | -0.0109 | -0.0827; 0.0608 | 0.765 |
| **3. Fatigue** |  |  |  |  |
|  | Mild [1] | -0.0061 | -0.1067; 0.0945 | 0.905 |
|  | Moderate [2] | 0.0032 | -0.0903; 0.0966 | 0.947 |
|  | Severe [3] | -0.0148 | -0.1106; 0.0811 | 0.762 |
| **4. Smell / taste** |  |  |  |  |
| Altered smell | Mild [1] | 0.0160 | -0.0284; 0.0604 | 0.478 |
|  | Moderate [2] | 0.0594 | 0.0055; 0.1134 | 0.031 |
|  | Severe [3] | 0.0451 | -0.0248; 0.1150 | 0.205 |
| Altered taste | Mild [1] | -0.0145 | -0.0568; 0.0278 | 0.502 |
|  | Moderate [2] | -0.0627 | -0.1146; -0.0108 | 0.018 |
|  | Severe [3] | -0.0683 | -0.1398; 0.0031 | 0.061 |
| **5. Pain / discomfort** |  |  |  |  |
| Chest pain | Mild [1] | -0.0161 | -0.0478; 0.0155 | 0.318 |
|  | Moderate [2] | 0.0188 | -0.0167; 0.0543 | 0.299 |
|  | Severe [3] | -0.0053 | -0.0656; 0.0550 | 0.863 |
| Joint pain | Mild [1] | -0.0130 | -0.0543; 0.0284 | 0.538 |
|  | Moderate [2] | -0.0209 | -0.0663; 0.0245 | 0.367 |
|  | Severe [3] | -0.0677 | -0.1225; -0.0129 | 0.016 |
| Muscle pain | Mild [1] | -0.0308 | -0.0739; 0.0124 | 0.162 |
|  | Moderate [2] | -0.0306 | -0.0775; 0.0163 | 0.200 |
|  | Severe [3] | -0.0600 | -0.1175; -0.0024 | 0.041 |
| Headache | Mild [1] | -0.0125 | -0.0476; 0.0226 | 0.486 |
|  | Moderate [2] | -0.0135 | -0.0495; 0.0225 | 0.461 |
|  | Severe [3] | 0.0148 | -0.0324; 0.0620 | 0.539 |
| Abdominal pain | Mild [1] | -0.0053 | -0.0374; 0.0268 | 0.745 |
|  | Moderate [2] | -0.0069 | -0.0450; 0.0311 | 0.721 |
|  | Severe [3] | 0.0099 | -0.0465; 0.0663 | 0.731 |
| **6. Cognition** |  |  |  |  |
| Problems with concentration | Mild [1] | -0.0064 | -0.0646; 0.0517 | 0.828 |
|  | Moderate [2] | 0.0163 | -0.0448; 0.0773 | 0.601 |
|  | Severe [3] | 0.0217 | -0.0492; 0.0926 | 0.548 |
| Problems with memory | Mild [1] | -0.0109 | -0.0611; 0.0393 | 0.670 |
|  | Moderate [2] | -0.0174 | -0.0719; 0.0370 | 0.530 |
|  | Severe [3] | -0.0289 | -0.0939; 0.0361 | 0.383 |
| Problems with planning | Mild [1] | 0.0210 | -0.0194; 0.0615 | 0.308 |
|  | Moderate [2] | -0.0078 | -0.0528; 0.0372 | 0.734 |
|  | Severe [3] | -0.0061 | -0.0661; 0.0540 | 0.842 |
| **7. Palpitations / dizziness** |  |  |  |  |
| Palpitations in certain positions, activity, rest | Mild [1] | 0.0026 | -0.0312; 0.0364 | 0.879 |
|  | Moderate [2] | 0.0069 | -0.0293; 0.0432 | 0.707 |
|  | Severe [3] | 0.0181 | -0.0359; 0.0720 | 0.511 |
| Dizziness in certain positions, activity, rest | Mild [1] | -0.0051 | -0.0401; 0.0300 | 0.777 |
|  | Moderate [2] | 0.0008 | -0.0360; 0.0376 | 0.965 |
|  | Severe [3] | -0.0133 | -0.0620; 0.0355 | 0.594 |
| **8. Post-exertional malaise** |  |  |  |  |
| Crashing or relapse after physical, cognitive or emotion exertion | Mild [1] | -0.0307 | -0.0820; 0.0207 | 0.241 |
|  | Moderate [2] | -0.0069 | -0.0545; 0.0407 | 0.775 |
|  | Severe [3] | 0.0066 | -0.0461; 0.0592 | 0.807 |
| **9. Anxiety / mood** |  |  |  |  |
| Feeling anxious | Mild [1] | -0.0140 | -0.0604; 0.0325 | 0.555 |
|  | Moderate [2] | -0.0448 | -0.0928; 0.0033 | 0.068 |
|  | Severe [3] | -0.0799 | -0.1378; -0.0221 | 0.007 |
| Feeling depressed | Mild [1] | -0.0332 | -0.0716; 0.0051 | 0.089 |
|  | Moderate [2] | -0.0327 | -0.0747; 0.0093 | 0.127 |
|  | Severe [3] | -0.1003 | -0.1530; -0.0477 | <0.001 |
| Having unwanted memories of your illness or time in hospital | Mild [1] | -0.0276 | -0.0662; 0.0109 | 0.160 |
|  | Moderate [2] | -0.0086 | -0.0610; 0.0439 | 0.748 |
|  | Severe [3] | 0.0174 | -0.0530; 0.0878 | 0.628 |
| Having unpleasant dreams about your illness or time in hospital | Mild [1] | 0.0051 | -0.0361; 0.0462 | 0.809 |
|  | Moderate [2] | -0.0269 | -0.0819; 0.0281 | 0.338 |
|  | Severe [3] | -0.0285 | -0.1133; 0.0563 | 0.509 |
| Trying to avoid thoughts or feelings about your illness or time in hospital | Mild [1] | -0.0236 | -0.0583; 0.0111 | 0.182 |
|  | Moderate [2] | -0.0195 | -0.0674; 0.0285 | 0.425 |
|  | Severe [3] | -0.0154 | -0.0862; 0.0554 | 0.670 |
| **10. Sleep problems** |  |  |  |  |
|  | Mild [1] | -0.0132 | -0.0651; 0.0387 | 0.619 |
|  | Moderate [2] | -0.0218 | -0.0714; 0.0277 | 0.387 |
|  | Severe [3] | -0.0401 | -0.0921; 0.0120 | 0.131 |
| **11. Communication** |  |  |  |  |
|  | Mild [1] | 0.0035 | -0.0314; 0.0385 | 0.842 |
|  | Moderate [2] | 0.0228 | -0.0173; 0.0630 | 0.265 |
|  | Severe [3] | 0.0270 | -0.0272; 0.0813 | 0.328 |
| **12. Walking or moving around** |  |  |  |  |
|  | Mild [1] | -0.0387 | -0.0758; -0.0016 | **0.041** |
|  | Moderate [2] | -0.0849 | -0.1257; -0.0441 | **<0.001** |
|  | Severe [3] | -0.1470 | -0.2003; -0.0938 | **<0.001** |
| **13. Personal care** |  |  |  |  |
|  | Mild [1] | -0.0739 | -0.1084; -0.0394 | **<0.001** |
|  | Moderate [2] | -0.1063 | -0.1518; -0.0609 | **<0.001** |
|  | Severe [3] | -0.2014 | -0.2671; -0.1356 | **<0.001** |
| **14. Other activities of daily living** |  |  |  |  |
|  | Mild [1] | -0.0506 | -0.1081; 0.0069 | 0.084 |
|  | Moderate [2] | -0.0565 | -0.1134; 0.0004 | 0.051 |
|  | Severe [3] | -0.0715 | -0.1358; -0.0072 | **0.029** |
| **15. Social role** |  |  |  |  |
|  | Mild [1] | -0.0318 | -0.0701; 0.0065 | 0.104 |
|  | Moderate [2] | -0.0206 | -0.0598; 0.0186 | 0.303 |
|  | Severe [3] | -0.0807 | -0.1317; -0.0297 | **0.002** |
| **Overall Health (0 – 10)** |  | 0.0154 | 0.0076; 0.0233 | **<0.001** |
| Age |  | 0.0009 | -0.0000; 0.0018 | 0.057 |
| Ethnicity [Black] |  | 0.0274 | -0.0671; 0.1219 | 0.569 |
| Ethnicity [Mixed] |  | -0.0128 | -0.1084; 0.0829 | 0.793 |
| Ethnicity [None] |  | -0.0384 | -0.1291; 0.0523 | 0.406 |
| Ethnicity [Other] |  | 0.1369 | -0.0057; 0.2796 | 0.060 |
| Ethnicity [White] |  | -0.0131 | -0.0634; 0.0372 | 0.609 |
| Sex [Male] |  | -0.0152 | -0.0418; 0.0115 | 0.264 |
| Pre-Long COVID Comorbidities |  | -0.0052 | -0.0238; 0.0134 | 0.584 |
| Observations |  | 737 |  |  |
| R<sup>2</sup> / R<sup>2</sup> adjusted |  | 0.71 / 0.67 |  |  |
| AIC |  | -585.2 |  |  |
| log-Likelihood |  | 396.6 |  |  |

**Supplementary Figure 1.**
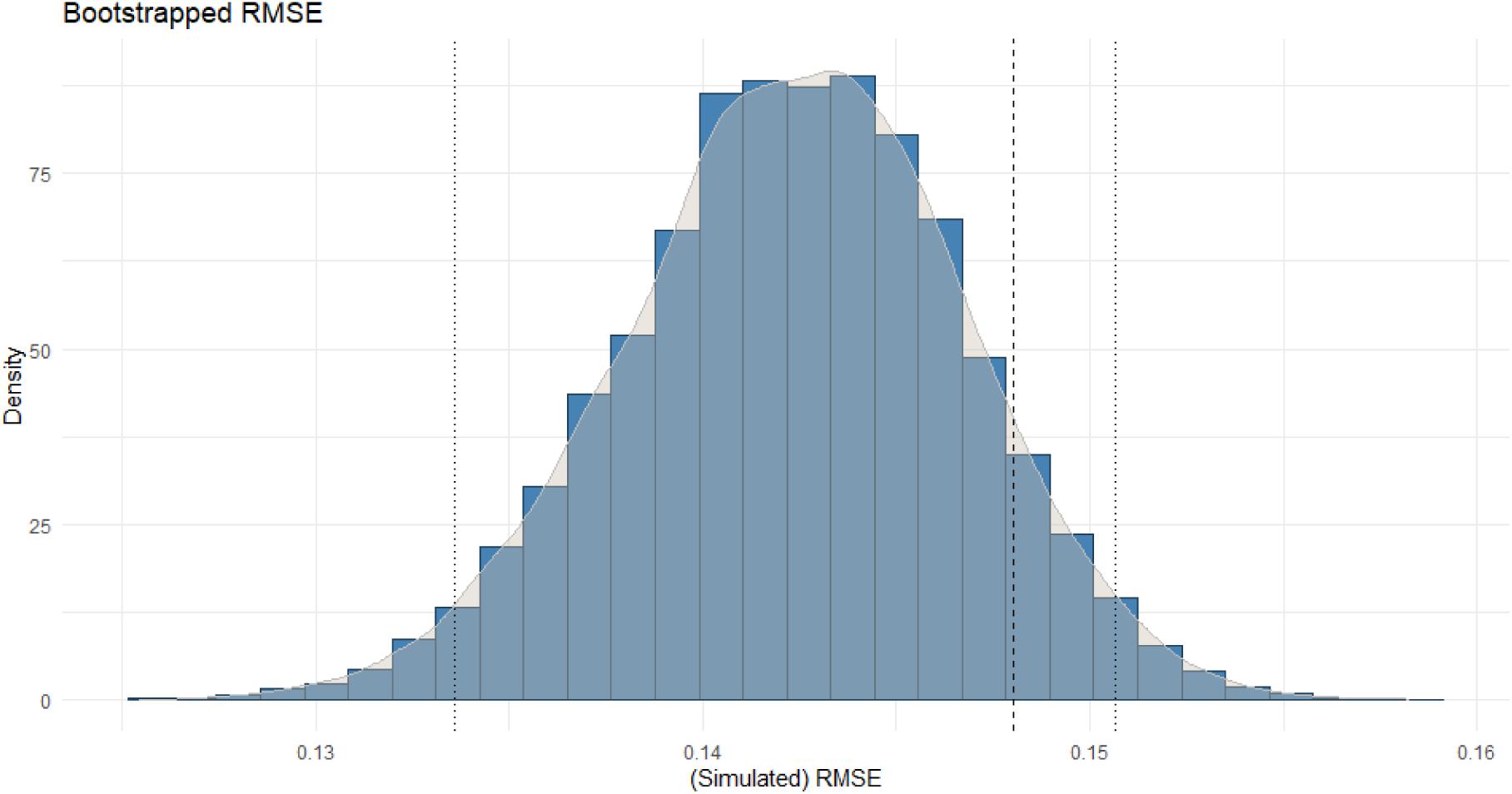
Bootstrapped RMSE.

